# A randomised clinical trial of STAtin therapy for Reducing Events in the Elderly (STAREE): Statistical analysis plan

**DOI:** 10.1101/2025.02.24.25321974

**Authors:** Rory Wolfe, Stephane Heritier, Ella Zomer, Mary Lou Chatterton, Andrea Curtis, Simone Spark, Zachary Flanagan, Trever T-J Chong, Geoffrey C Cloud, Mark R Nelson, Stephen J Nicholls, Chris Moran, Christopher M Reid, Sophia Zoungas, the STAREE Investigator Group

**Affiliations:** School of Public Health and Preventive Medicine, Monash University, Melbourne, Victoria, Australia; Monash University Clinical Trials Centre, Monash University, Melbourne, Victoria, Australia; School of Psychological Sciences & Turner Institute of Brain and Mental Health, Monash University, Melbourne, Victoria, Australia; Department of Neuroscience, School of Translational Medicine, Monash University, Melbourne, Victoria, Australia; Menzies Institute for Medical Research, University of Tasmania, Hobart, Tasmania, Australia; Victorian Heart Institute, Monash University, Clayton, Victoria, Australia; School of Population Health, Curtin University, Perth, Western Australia, Australia

## Abstract

The STAREE randomised controlled trial of 9971 community-dwelling older adults without cardiovascular disease, diabetes or dementia is designed to compare daily 40mg atorvastatin to placebo. We present a statistical analysis plan to pre-specify the analytical approaches that will be taken when reporting the main findings from the trial. The plan describes the analysis approach for the co-primary endpoints: (a) death, dementia, and persistent physical disability; (b) major cardiovascular events; and all secondary endpoints. A pre-specified economic evaluation is also included as part of this plan.

## 1 INTRODUCTION

### 1.1 Summary

This Statistical Analysis Plan (SAP) provides a detailed statement of the intended statistical analyses, including the economic evaluation, to be performed on data collected in the STAtin therapy for Reducing Events in the Elderly (STAREE) trial in order to address the study’s main hypotheses.

The SAP draws extensively from the study protocol (Version 3.2, May 2024 and Version 3.2b, November 2025) and the published protocol paper (Zoungas 2023), with the SAP providing further detail on the analytical methods and relevant endpoint definitions. Extracts from the protocol (or protocol paper) are presented in *blue italicised font*. Details on randomisation procedures and sample size are not included here and the reader is referred to the protocol.

The SAP can undergo revision outside of the protocol. It is not anticipated that revisions of the SAP would require review by an ethics committee. No changes to the SAP will be made after database lock or any unblinding of study investigators.

The SAP complies with International Conference on Harmonisation (ICH) E9 Guidelines including ICH E9(R1) for handling intercurrent events such as competing risks, loss to follow-up and compliance with study drug over the duration of follow-up (International Conference on Harmonisation 2020).

### 1.2 Background

*STAREE is a double blind, randomised placebo-controlled parallel group trial that is investigating whether statin therapy can prolong good health and maintain independence amongst older people*.

The study will enrol men and women 70 years of age and over who do not have a history of clinical cardiovascular disease events, diabetes or dementia and live independently in the community.

The trial’s primary objectives are to *determine in people aged* ≥*70 years the effect of statin therapy (40 mg atorvastatin) versus placebo on two co-primary clinical endpoints;*

i. *Disability-free survival defined as survival free of dementia and persistent physical disability (a composite of all cause death or development of dementia or persistent physical disability); and*
ii. *Major cardiovascular events (a composite of cardiovascular death, non-fatal myocardial infarction, non-fatal stroke* or coronary revascularisation).

### 1.3 Null hypotheses

1. Treatment with atorvastatin 40mg compared with placebo over an average 6-year period will not prolong overall disability-free survival among a primary prevention population aged ≥70 years.
2. Treatment with atorvastatin 40mg compared with placebo over an average 6-year period will not reduce the incidence of major cardiovascular events among a primary prevention population aged ≥70 years.

### 1.4 Alternative hypotheses

1. Treatment with atorvastatin 40mg over an average 6-year period will prolong overall disability-free survival among a primary prevention population aged ≥70 years.
2. Treatment with atorvastatin 40mg over an average 6-year period will reduce the incidence of major cardiovascular events among a primary prevention population aged ≥70 years.

## 2 ENDPOINTS

Detailed definitions of the primary and secondary endpoints and other measures, and the endpoint ascertainment process and adjudication are provided in the protocol and its Appendix 2.

### 2.1 Co-primary composite endpoints

STAREE will have two co-primary endpoints, disability free-survival and major cardiovascular events, which are both composite endpoints.

1. *Disability-free survival defined as survival free of dementia and persistent physical disability (a composite of all cause death or development of dementia or persistent physical disability); and*
2. *Major cardiovascular events defined as the first occurrence of cardiovascular death, non-fatal myocardial infarction, non-fatal stroke* or coronary revascularisation.

A change to the composite major cardiovascular event endpoint was made after publication of the trial protocol paper (Zoungas 2023). The original “3-point” composite definition only included cardiovascular death, non-fatal myocardial infarction and non-fatal stroke. After an average participant follow up of 3 years (in Q1, 2024), the observed event rates among all participants were reviewed (blinded to randomisation and treatment) and noted to be much lower than anticipated for this “3-point” major cardiovascular events endpoint. To maintain the power of the study and achieve the required number of events at completion of the trial (anticipated in Q4, 2025), the trial steering committee decided to add coronary revascularisation to this co-primary composite endpoint. The previous “3-point” composite definition is now listed as a pre-specified secondary endpoint.

Coronary revascularisation is defined as any arterial revascularisation event which is determined by an expert clinical endpoint adjudication committee (EAC) as involving a major epicardial coronary artery.

### 2.2 Secondary endpoints

*Two types of outcome measures are pre-specified secondary endpoints in STAREE. The first group are components of the co-primary endpoints and additional cognitive and cardiovascular endpoints (heart failure and all revascularisation procedures). The second group include non-cardiovascular endpoints (need for permanent residential care, hospitalisation and cancer)*.

1. Major cardiovascular events defined as the first occurrence of cardiovascular death, non-fatal myocardial infarction or non-fatal stroke.
2. *All cause death*
3. *Cardiovascular death*
4. *Fatal and non-fatal myocardial infarction*
5. *Fatal and non-fatal stroke*
6. *Coronary revascularisation*
7. *Persistent physical disability (in* Activities of Daily Living *based on Life Ability questionnaire)*
8. *Dementia*
9. *Other cognitive impairment (not meeting criteria for Dementia)*
10. *Approved need for permanent residential care (based on reporting by an Aged Care Assessment Team)*
11. *All cause hospitalisation (reasons and length of stay)*
12. *Heart failure*
13. *Atrial fibrillation*
14. *All revascularisation procedures*
15. *Fatal and non-fatal cancer (excluding non-melanoma skin cancer)*
16. *Quality of life (SF-36 questionnaire)*
17. *Cost-effectiveness*

### 2.3 Tertiary objectives

*To determine the effects of statin therapy (atorvastatin 40 mg) versus placebo on each of fasting glucose/glycated haemoglobin, urine albumin to creatinine ratio (*urine *ACR), estimated glomerular filtration rate (eGFR), frailty phenotype, loss of functional independence (incident instrumental activities of daily living disability), and depressive symptoms (incident and recurrent)*.

As some participants may have depressive symptoms at baseline (a history of depression or a Centre for Epidemiologic Studies Depression Scale (CES-D)-10 score of 10 or above), both pooled and separated analyses of incident depressive symptoms will be undertaken (CES-D-10 at any post-randomisation data collection) in those symptom-free at baseline, and recurrent depressive symptoms in those with symptoms at baseline.

### 2.4 Estimands

For all estimands of interest (International Conference on Harmonisation 2020) the following apply in each case:

**Population-level summary measure**:average treatment effect unless otherwise specified.

**Endpoint** is time from randomisation until the first occurrence of one of the primary or secondary endpoints.

**Target population** is independent community-dwelling adults aged 70 years and older free of any history of cardiovascular disease events, dementia or diabetes, attending primary care with their general practitioner.

**Treatment condition** is daily 2x20mg atorvastatin tablets for an average 6-year period and compared to identical placebo, including an initial 4-week tolerability assessment of one tablet per day followed by increase to two tablets if tolerated. This includes allowance for subsequent dose reduction to one 20mg atorvastatin tablet per day or temporary interruption to manage tolerability, and allowance for commencement of open-label lipid-lowering treatment according to treating clinician advice in response to a participant experiencing a recognised clinical indication (such as a myocardial infarction or stroke).

**Intercurrent events and strategies to address these**: (i) the occurrence of death (for non-fatal endpoints) and certain causes of death (for endpoints that include specific causes of death) will be treated as competing risks; (ii) the withdrawal of consent is anticipated in only a small number of participants and such individuals will be analysed on the assumption of withdrawal at random relative to the outcome events being analysed; (iii) the cessation of statin use (for randomised statin group) or commencement of prescribed statins (for randomised placebo group) will be dealt with in different ways depending on the treatment effect to be estimated.

## 3 DATA ANALYSIS

### 3.1 Interim analysis

*One pre-specified interim analysis may occur when an average of 3.25 years of follow-up per participant has accrued and >50% of the primary endpoints have occurred. The DSMB will be responsible for reviewing any interim analysis on unblinded data and providing recommendations to the trial executive committee. The DSMB will review deaths, serious adverse events and other endpoint data on a periodic basis*.

The stopping rule will be if the death, dementia, persistent physical disability endpoint achieves significance with positive effects on the outcome at the interim analysis then the DSMB may recommend cessation for benefit. The p-value boundary for comparison of treatment groups at this interim analysis will be 0.001.

### 3.2 Final analysis

*The data will be analysed by statisticians based at the Biostatistics Unit*, School of Public Health and Preventive Medicine, *Monash University*.

The analyses will be performed using the statistical software packages Stata and R.

*Baseline characteristics of the two treatment groups will be tabulated* using mean and standard deviations for continuous measures that follow a reasonably symmetrical distribution on their scale of measurement or after logarithm transformation, or medians and interquartile ranges otherwise, and numbers and percentages for categorical measures. No p-values will be calculated for this table since a null hypothesis of no difference between the two groups at baseline (i.e. that the two groups are random samples from the same underlying population) is known to be true due to the randomisation process that is used in the trial as specified in the protocol. Imbalance will be *defined as a 0.25 standard deviation difference in means (quantitative characteristics) or an odds ratio of 1.5 (binary characteristics)*.

Safety will be assessed with tabulation of events known to be possible side-effects of statin use: new diagnosis of diabetes, new diagnosis of myopathy (muscle symptoms), liver impairment, and venous thromboembolism, each as defined in the Protocol. Serious adverse events will similarly be tabulated. These tabulations will be stratified by randomised treatment and will be performed for the full ITT set and the modified ITT set.

*All primary comparisons between treatment arms will be on an intention-to-treat basis, that is, according to the group to which participants were randomised and without reference to their actual compliance with assigned treatment*. Participants who did not take any study drug (regardless of randomised group) will be excluded in a secondary analysis, i.e. a modified intention to treat approach will be taken for a sensitivity analysis.

*Each of the co-primary endpoints will be analysed separately in time-to-event analyses. Event rates (time to first event within each endpoint definition) will be compared between groups using a hazard ratio, 95% confidence interval and two-sided p-value from a Cox proportional hazards regression model fitted to the endpoint with censoring for individuals not experiencing an endpoint event at their most recent study visit, and a single covariate being an indicator of the group to which the individual was randomised, statin or placebo. Given the large sample size, randomisation is anticipated to adequately balance baseline characteristics of participants in the two treatment groups. Hence unadjusted analyses will be considered primary*.

For the co-primary endpoints, the time to event will be defined as follows. For disability-free survival, the time from randomisation date to the earliest of: (a) date of death; or (b) date of dementia defined as the date at which both instrumental ADL assessment and cognitive test results are available to confirm dementia diagnosis or, in the case of adjudicated dementia events reported from participant/carer/doctor/death certificate/admission to care/cognitive test triggers, the date of dementia diagnosis (if this is unknown then an adjudicated date of diagnosis is used); or (c) date of persistent physical disability defined as date of ADL administration at which physical disability was identified and subsequently confirmed 6 months later as being persistent, or adjudicated date for events detected from other sources and confirmed by adjudication. For major cardiovascular events, the time to event is defined as time from randomisation date to the date of occurrence of an adjudicated endpoint.

For participants who do not experience a primary endpoint, the administrative censoring date will be: (i) for disability-free survival, the end date of the trial’s intervention phase; (ii) for major cardiovascular events, the earliest of date of death, loss to follow up, or the end date of the trial’s intervention phase. *Loss to follow-up will be considered a censoring event. This equates to an assumption that data is missing at random given the participant’s treatment group and the timing of their loss to follow-up*. One exception will be for the endpoint all-cause death (and endpoints that include all-cause death where death constitutes most of the endpoint events) because loss to follow up does not preclude capture of fact of death from public records which the study regularly checks.

In analyses of major cardiovascular events, death due to causes other than those specified by the endpoint will be considered as censoring events, i.e. cause-specific hazard ratios will be estimated, but for graphical representation these will be treated as competing risks and cumulative incidence plots will be shown with cumulative incidence estimated using the Aalen–Johansen estimator (Aalen 1978).

*A closed testing procedure will be used to allow for the multiple testing arising from two co-primary endpoints. This approach is based on the expectation that cardiovascular benefit will be the main contributor to improved disability-free survival and that a substantial effect of statins on the latter is unlikely in the absence of an effect on the former. First, major cardiovascular events will be tested at alpha=0.05 and, if the major cardiovascular events p value is <0.05 then second, disability-free survival will be tested at alpha=0.05. If the major cardiovascular events p value is not <0.05 then a p value for disability-free survival will not be presented*.

For composite endpoints we will also undertake total events analyses, similarly to previous cardiovascular trials (e.g. Bhatt 2019) and with allowance for re-occurrences of the same event within the endpoint definition as well as occurrence of other events. In this respect, secondary *analyses of the co-primary endpoints will be performed using* the Andersen-Gill *extended Cox proportional hazards regression model based on time(s) to any event within the endpoint definition (for relevant endpoints that can occur more than once)*. The Andersen-Gill approach provides enhanced power, straightforward generalisation to covariate adjustment through its regression model structure, simple assumptions regarding baseline hazards and treatment effect, and has been shown to be a reasonable choice in composite endpoint scenarios (Ozga 2018, Marceau West 2024). In sensitivity analyses, other model structures for recurrent events will be examined, a multistate model will be used to analyse occurrences of events of different types within an endpoint definition, and bundling will be used whereby non-fatal events, occurring within a narrow timeframe prior to a death due to a corresponding cause, will be omitted. Finally, a novel composite endpoint comprised of all events from the two co-primary endpoints will also undergo a total events analysis.

Secondary endpoints in the form of time to event will be analysed using univariable Cox proportional hazards regression models. Censoring times for secondary endpoints will be defined as for major cardiovascular events except for all-cause death which will be defined as for disability-free survival. All confidence intervals (CI) will be reported as 95% CI. In analyses of these secondary endpoints, death due to causes other than those specified by the endpoint will be considered as censoring events.

The secondary endpoint persistent physical disability will be analysed with an illness death model and censoring will be at the time of the event or, in the absence of an event, at the second last visit as this censoring option has been demonstrated to be less biased than other approaches (Thao 2024). For time-to-event analyses for endpoints that include events for which the exact date of occurrence is unknown and based on screening at specific regular intervals as outlined in the protocol, for example persistent physical disability from ADLs, sensitivity analyses will be conducted using interval-censored proportional hazards regression models.

The secondary endpoint all fatal and non-fatal stroke will be compared between treatment groups using Cox proportional hazards time to first event models. Additional secondary analyses of this endpoint will include separate analyses of:

i. ischaemic stroke – overall, confirmed by TOAST criteria (Adams 1993), and by subcategories of large artery atherosclerosis, cardioembolism, small artery occlusion/disease, tandem, other aetiology (including CVST, systemic illness, etc), undetermined (inconclusive/incomplete testing available), haemorrhagic infarct, embolic stroke of undetermined source, and
ii. haemorrhagic stroke – overall and by subcategories of deep [basal ganglia/brainstem], lobar, ventricular (primary/secondary), and spontaneous subarachnoid haemorrhagic stroke (aneurysmal/non-aneurysmal/undetermined).

The secondary endpoint myocardial infarction will be compared between treatment groups using Cox proportional hazards time to first event models including separate analyses of type 1 (atherothrombotic coronary artery disease) and type 2 (oxygen supply and demand imbalance) myocardial infarction (Thygesen 2018).

The secondary endpoints coronary revascularisation and all revascularisation procedures will be compared between treatment groups using Cox proportional hazards time to first event models including separate analyses of emergency and planned revascularisation procedures.

The secondary endpoint quality of life will be measured by the SF-36 Physical Component Summary (PCS) and Mental Component Summary (MCS) scores. The PCS and MCS will be analysed in linear mixed effect models that include random intercept and slope effects for individuals and fixed effects for randomised treatment group, time, and an interaction of treatment group and time (De Livera 2014). The interaction p-value from these models will be used to determine whether, in the context of primary prevention, statin impacts the trajectory of quality of life in older age differently from placebo. Finally, a separate Health Economic Analysis Plan for the STAREE trial is included below to pre-specify the methods for cost effectiveness.

To address tertiary objectives, we will employ linear mixed effect models for continuous outcomes that include random intercept and slope effects for individuals and fixed effects for randomised treatment group, time, and an interaction of treatment group and time (De Livera 2014). The interaction term from these models will be used to assess the effects of statin therapy on trajectories of pathology measures. Tertiary measures such as urine ACR will be log transformed before analysis if they follow a right-skew distribution rather than a symmetric distribution. Reaching the frailty phenotype and losing functional independence will be analysed using interval-censored proportional hazards regression models. Incident and recurrent depressive symptoms will be analysed using binary logistic regression for the first post-randomisation CES-D-10 measure with an interaction between visit number at which that measure occurred (year 2, 3 etc) and randomised treatment group.

*No statistical adjustment will be made for the multiple secondary endpoints and no p values will be presented for the statin versus placebo comparison for these endpoints. The reporting of all secondary endpoint analyses will make clear whether either of the co-primary endpoints were statistically significant*. The rationale for this is that for these analyses type 2 error is of greater concern and this concern would be worsened if we reduce type 1 error through multiplicity adjustment.

*The proportional hazards assumption will be tested for each model*. The proportional hazards assumption will be tested in each of the main analyses and a p-value for the test will be presented – however due to the possibility of large numbers of events for several of the outcomes and excessive power for this test of interaction, its p-value will be discussed together with an interpretation of the extent to which a plot of log(-log) transformed estimated survival probabilities against log-transformed at-risk time demonstrates parallel lines for the two treatment groups. If based on this discussion the proportional hazards assumption is considered inappropriate, then a sensitivity analysis will be undertaken to examine the evidence for a time-dependent effect of statin.

Difference in restricted mean survival time (RMST) or restricted mean time lost (RMTL – if censoring due to death or competing causes of death is relevant) between randomised groups will be calculated as a treatment effect to be reported, either alongside the corresponding hazard ratio or, in cases where the proportional hazards assumption is found to be violated, in preference to an overall hazard ratio.

Some composite endpoints include a mix of events for which exact date of occurrence is known in some type(s) of event (such as all-cause death) and not known in other type(s) of event (such as physical disability persistence from ADLs). In this situation, sensitivity analyses will be conducted using time-to-event methods that allow for component-wise censoring (Eaton 2024).

A set of secondary analyses will be undertaken to analyse participants and follow-up time according to whether study drug was being taken or not (see Section 3.5.2).

Analyses that are undertaken and reported but which are not included in this SAP will be labelled in all reports as not being pre-specified (or post-hoc).

### 3.3 Sub-group analyses

Analyses will be undertaken within sub-groups of participants for whom there may be different effects of statin. The p-values for interaction terms in multivariable Cox proportional hazards regression models will be used to test for heterogeneity of treatment effect of atorvastatin between sub-groups. Subgroup analyses will report effect estimates and confidence intervals for each subgroup along with the p-value for the test of interaction.

The pre-specified list of sub-groups is:

a. **Males versus females**
b. **Age** ≥**75 versus <75 years:** The mean age of participants at randomisation was 75 years.
c. **Baseline blood pressure level:** Comparisons among tertiles of each of systolic and diastolic blood pressure.
d. **Body mass index:** defined according to World Health Organization criteria as normal weight (<25 kg/m^2^), overweight (25-29.9 kg/m^2^) and obese (≥30 kg/m^2^).
e. **Smoking status:** Never, Past or Current smokers.
f. **Baseline 3MS score**: Two groups defined by split at median 3MS score (below median versus at or above median score).
g. **eGFR**: <60 v ≥60 mL/min per 1.73m^2^ calculated using the Chronic Kidney Disease Epidemiology Collaboration 2021 equation using serum creatinine.
h. **Baseline cholesterol level**: Comparisons among tertiles of each of LDL-C and HDL-C.
i. **Period of recruitment relative to COVID-19 pandemic in Australia**: Two groups split according to date of randomisation being before or after February 2020.

### 3.4 Additional Sensitivity analyses

In time to event analyses, treating loss to follow-up as a censoring event equates to an assumption that data is missing at random given the participant’s treatment group and the timing of their loss to follow-up. *The adequacy of this assumption will be checked in sensitivity analyses that will include both imputation approaches and adjustment for baseline covariates predictive of propensity for dropout*. Where appropriate, imputation using retrieved dropouts will be considered as has been recently recommended (Medcalf 2026).

A secondary set of analyses will be performed to adjust for any baseline characteristics that are found to be imbalanced between groups according to the definition in Section 3.2. These analyses will be conducted using multivariable Cox proportional hazards regression models. Age, sex and comorbidities (at baseline) that are thought to be strongly prognostic of outcome in this population, will be included in these adjusted analyses.

*The impacts of COVID-19 will be investigated in sensitivity analyses that include examination of participant event rates by whether their randomisation occurred pre-pandemic, peri-pandemic or post-pandemic, and consider COVID-19 vaccination status (which will be an intercurrent event for those randomised pre-pandemic and a baseline covariate for those randomised post-pandemic), following emerging guidance on analytical approaches and estimand definitions (van Lancker 2023)*. This will include a pre-specified subgroup analysis by COVID-19 period.

### 3.5 Analysis populations

#### 3.5.1 Intention to treat (ITT) set

The ITT set will include all participants randomised, including participants who were identified after randomisation as not satisfying all inclusion criteria at the time of randomisation. A modified ITT set will be the ITT set excluding participants who did not take any study drug (regardless of randomised group). A second analysis will censor at-risk time when unblinding related to an individual study participant occurred, and with exclusion of any participants in the modified ITT set that were identified, after randomisation, to not meet study inclusion and exclusion criteria.

#### 3.5.2 Per-protocol analyses

A compliance adjusted analysis will be undertaken by applying inverse probability of censoring weighting at the time of treatment switches, with a switch being identified from changed satisfaction of the treatment condition specified for estimands in Section 2.5. Follow-up will be discretised into intervals such as 30-days, and pooled logistic regression will be used for both the weight-generating and outcome models. The weight-generating model will be stratified by randomised treatment group. The weights will be stabilised and truncated, for example at the 99.9^th^ (and 0.1^st^) percentiles. Robust standard errors will be used in the outcome model to account for within-individual correlation across intervals.

### 3.6 Sub-studies

Additional cognitive outcomes on a subset of participants will be analysed for the STAREE-MIND substudy as detailed elsewhere (Harding 2023). Similarly, additional cardiac measures on a subset of participants will be analysed for the STAREE-HEART sub-study (Hopper 2024).

### 3.7 Economic evaluation

#### 3.7.1 Overview

The primary objective of the health economic evaluation is to calculate the cost-effectiveness of atorvastatin therapy versus placebo (no atorvastatin therapy) over the trial duration using a within-trial economic evaluation study design. The second objective is to build an economic model to extrapolate the longer-term costs and benefits of atorvastatin therapy beyond the trial period over a lifetime for the Australian population.

A perspective of the government as payer will be adopted in the primary analysis and will include costs borne by the Commonwealth and State governments. A broader health sector perspective including both government costs and consumer out of pocket payments will be used in a secondary analysis.

In Australia, economic evaluations typically use the outcome metric of quality adjusted life years (QALYs) gained because cost-effectiveness ratios using QALYs have inherent value-for-money connotations. While there is no formal willingness to pay threshold in Australia, we will employ the most commonly used and accepted threshold of $50,000 per QALY gained (Carter 2008). The threshold will be varied in scenario analyses, as there is additional evidence suggesting a lower threshold of around $28,000 per QALY gained (Edney 2018).

Costs and outcomes will be presented as undiscounted and discounted at 5% as per Australian Government guidelines (Commonwealth of Australia Department of Health 2016). The discount rate will be varied to 1.5% in scenario analyses as per recent recommendations (Medicines Australia 2022).

Consolidated Health Economic Evaluation Reporting Standards (CHEERS) 2022 guidelines (Husereau 2022) and ISPOR best practice modelling guidelines (Caro 2012) will be followed when reporting the health economic evaluation, and in a format appropriate to stakeholders (e.g. PBAC, consumers, and national peak bodies) and policy makers.

#### 3.7.2 Costs

The costs to the government include the cost of atorvastatin therapy as well as other health care services utilised by study participants during the study period. All healthcare use will be valued in monetary terms using appropriate Australian unit costs in Australian dollars estimated at the time of analysis and presented, and valued in Australian dollars (AU$) for the most recent reference year (e.g., 2026 or 2027).

For the primary analysis, health service use will be derived from trial data. General practitioner consultations will be costed using a weighted average cost paid by the government, derived from the Medicare Benefits Schedule (MBS) item reports (Department of Human Services 2019). Hospitalisations will be ascertained through trial records, and valued using Australian-Refined Diagnosis Related Groups (AR-DRGs), National Weighted Activity Unit (NWAU; Independent Hospital Pricing Authority 2018) and the most recently available published National Efficient Price. These cost weights underlie Activity Based Funding models for hospital care in Australia (Independent Hospital Pricing Authority 2018). Adjustments will be made for inflation as necessary. Medication use will be determined through trial records, and valued using Pharmaceutical Benefits Scheme (PBS) item prices (Department of Health and Aged Care 2025).

A secondary analysis employing health service use from linked national and state/territory government administrative datasets, using a broader health sector perspective, which includes consumer out of pocket costs, will be conducted. This will include MBS claims which include all health professional consultations (GPs, specialists, etc), pathology, and imaging procedures. The PBS claims include all government reimbursed prescription medications. Both of these administrative data sources include information on the cost paid by the government and the out-of-pocket cost to consumers.

The average per patient healthcare use in each year over the trial duration will be tabulated by randomised treatment group, with precision estimates (i.e., standard deviations, SD and 95% CIs). The difference in healthcare use between the intervention and control group will be presented by category (e.g., medical consultations, medication use, etc) with 95% CIs. The total cost for each participant will be calculated as the sum of atorvastatin cost and all concurrent healthcare costs over the study period.

#### 3.7.3 Outcomes

The primary outcome measure will be QALYs. Utility values will be derived from participant responses at each time point on the 36-Item Short Form Survey Instrument (SF-36) quality of life instrument using the SF-6D preference-based scoring algorithm (Brazier 2004).

The utility values recorded at baseline and at each follow-up over the trial duration will be used to calculate total QALYs for each participant using the area under the curve method (Glick 2014), adjusting for any imbalances in baseline SF-6D values.

Disability-free survival and major cardiovascular events prevented will also be examined as additional outcome measures

#### 3.7.4 Statistical analysis of economic data

Mean differences in costs, QALYs and net benefits between the randomised treatment groups will be estimated with associated 95% CIs. All statistical tests will be two-sided, and the statistical significance level will be set at < 5%.

The statistical analyses for the economic evaluation will follow the principles detailed previously for the primary analysis and will employ an ITT approach, where all individuals randomised will be included in the analysis by their allocated trial arm status regardless of whether they received all, part or none of the intended treatments.

For the base case, generalised linear models (GLM) using a gamma family and log link will be used to estimate the difference in the total health sector costs between the atorvastatin and placebo groups over the trial period due to the typically right skewed distribution of cost data. The mean difference in QALYs will be estimated using GLM with the gaussian family and identity link with adjustment for the baseline covariates sex, age and utility.

Negative binomial regression will be used to examine the between group differences for the components of total cost including health care consultations, medications, and hospital visits.

In addition to reporting descriptive statistics and differences between treatment groups for costs and outcomes, incremental cost-effectiveness ratios (ICERs) will be calculated. ICERs will be calculated as the mean difference in total cost divided by the mean difference in outcome (i.e. QALYs) between the two trial arms. The CIs around ICERs will be calculated using a nonparametric bootstrap procedure, with 1,000 iterations to reflect sampling uncertainty. The bootstrapped ICERs and the CIs will be graphically represented on cost-effectiveness planes, with a confidence ellipse around the ICER. A cost-effectiveness plane is a plot of the 1,000 bootstrapped incremental costs and outcomes across four quadrants. The north-east quadrant represents the intervention (atorvastatin) costing more as well as conferring greater benefits than placebo. The south-east quadrant shows the proportion of iterations where atorvastatin costs less but incurs greater benefits than placebo (i.e., a “dominant” intervention), the north-west quadrant shows the proportion of iterations where atorvastatin incurs a cost but fewer benefits than placebo (i.e., a “dominated” intervention) and, lastly, the south-west quadrant shows the proportion of iterations whereby atorvastatin costs less and has fewer benefits than placebo.

#### 3.7.5 Sensitivity and subgroup analyses

Sensitivity analyses will be conducted using complete-cases only using GLM, with and without adjustment for covariates. Complete cases will be records with HRQoL data and resource use data observed over the trial duration. The results for complete cost and health outcome data (i.e., those with no missing data) as well as a strict per-protocol analysis of the data will be provided to identify the impact of missing data on the analysis and any sensitivity to protocol violations.

Analyses will be undertaken within sub-groups of participants for whom there may be different effects of atorvastatin as specified in Section 3.3.

#### 3.7.6 Modelled economic evaluation

The costs and outcomes data from the within trial evaluation will be used to estimate the population cost-effectiveness of statin therapy over a lifetime horizon using economic modelling techniques. This will be undertaken using the epidemiological literature to estimate longer term trajectories as well as resource use implications. More details regarding the modelled economic evaluation will be provided after the within trial economic evaluation has been completed. The modelling will only be undertaken if atorvastatin is found to be effective.

A budget impact analysis will be conducted using best practice methods (Sullivan 2014) to assess the potential financial impact of atorvastatin therapy in older adults to the Australian health care system, over a 5 year timeframe. An epidemiological approach will first estimate the number of older Australians who are eligible for statin therapy for primary prevention using population datasets.

The cost of atorvastatin therapy in each year, over 5 years will be calculated. Changes in health service utilisation (including due to the reduction in events if appropriate) will be estimated. Overall costs as a result of these changes will be calculated per year and aggregated over 5 years. Net financial costs will be presented and a description of the uncertainty around the estimates will be provided. These analyses will provide evidence to the Federal Government about affordability and sustainability.

## Data Availability

On completion of the trial, and after publication of the primary and secondary outcomes of the study, requests for access to de-identified data (to be provided through a secure online environment) may be submitted to the researchers located at the School of Public Health and Preventive Medicine, Monash University, Melbourne, Australia.

